# Mobility changes following COVID-19 stay-at-home policies varied by socioeconomic measures: An observational study in Ontario, Canada

**DOI:** 10.1101/2024.01.29.24301938

**Authors:** Siyi Wang, Linwei Wang, Stefan D Baral, Gary Moloney, Jaimie Johns, Carmen Huber, Jaydeep Mistry, Kamran Khan, Amrita Rao, Naveed Janjua, Tyler Williamson, Alan Katz, Huiting Ma, Mathieu Maheu-Giroux, Rafal Kustra, Sharmistha Mishra

**Affiliations:** MAP-Centre for Urban Health Solutions, St. Michael’s Hospital, Unity Health Toronto, Toronto, ON, Canada; Department of Epidemiology, Johns Hopkins School of Public Health, Baltimore, MD, United States; BlueDot, Toronto, ON, Canada; British Columbia Centre for Disease Control, Vancouver, BC, Canada; School of Population and Public Health, University of British Columbia, Vancouver, BC, Canada; Centre for Health Evaluation and Outcome Sciences, University of British Columbia, Vancouver, BC, Canada; Department of Community Health Sciences, University of Calgary, Calgary, AB, Canada; Centre for Health Informatics, University of Calgary, Calgary, AB, Canada; Departments of Community Health Sciences and Family Medicine, Manitoba Centre for Health Policy, Rady Faculty of Health Sciences, University of Manitoba, Winnipeg, MB, Canada; Department of Epidemiology and Biostatistics, School of Population and Global Health, McGill University, Montréal, QC, Canada; Division of Biostatistics, Dalla Lana School of Public Health, University of Toronto, Toronto, ON, Canada; Department of Statistical Sciences, University of Toronto, Toronto, ON, Canada; Division of Infectious Diseases, Department of Medicine, University of Toronto, Toronto, ON, Canada; Dalla Lana School of Public Health, University of Toronto, Toronto, ON, Canada; Institute of Health Policy, Management, and Evaluation, University of Toronto, Toronto, ON, Canada; Institute of Medical Sciences, University of Toronto, Toronto, ON, Canada

## Abstract

In Canada, lower income households and essential workers and were disproportionately at risk of SARS-CoV-2. Early in the pandemic, stay-at-home restriction policies were used to limit virus transmission. There remains an evidence gap in how changes in mobility, in response to the policies, varied across socioeconomic measures in Canada. The study objective was to describe the variability in mobility change to two restrictions, by neighborhood-level income and by proportion essential workers across five regions in Ontario, Canada. The first restriction was implemented on March 17, 2020 in all five regions; and the second restriction was implemented in November 23, 2020 in two of the regions. Using cell-phone mobility data aggregated to the census tract, we compared the average mobility (% of devices that travelled outside their “primary location”) three weeks before and after each restriction. We defined the adjusted mobility change via pre-restriction mobility subtracted from post-restriction, adjusted for 2019 levels. We used difference-in-differences analysis to quantify effect modification of the second restriction’s effect by socioeconomic measures. With the first restriction, crude mobility fell from 77.7% to 41.6% across the five regions. The adjusted mobility change to the first restriction was largest in the highest-income neighborhoods (-43.3% versus -38.4%) and in neighborhoods with the fewest essential workers (-44.5% versus -37.6%). The overall adjusted mobility change to the second restriction was small: -0.96% (95% confidence intervals, -1.53 to -0.38%). However, there was evidence of effect modification by socioeconomic measures (less pronounced decrease in lower-income neighborhoods and more essential workers). The findings suggest a temporal saturation effect of restrictions over subsequent waves, and a saturation effect by income and occupation, leading to prevention gaps across populations by socioeconomic measures. Findings highlight the need for tailored approaches at the intersections of income and occupation when addressing epidemics of novel and resurging respiratory pathogens.

## Introduction

In Canada, as within countries across the world, SARS-CoV-2 infections were disproportionately concentrated among people and communities experiencing social and economic marginalization [1]. The response during the first year of the pandemic centered on public health measures to reduce contact rates as a means to halt the spread of SARS-CoV-2. Measures included mandates to close non-essential business alongside limits on indoor gatherings and activities.

These restriction policies were met with early concerns about whom these policies could and could not reach [2, 3].

At the start of the pandemic, 60% of working-age adults in Canada were employed in jobs which could not be done remotely [4]. The front-facing jobs were also more likely to be lower-paid, classified as essential services during the pandemic, and included jobs in sales, trades, agriculture, manufacturing, transport, and the food industry [5]. The public health measures were designed as a universal policy, but individuals working in essential services would have to go to, and spend time at, their place of onsite work [1]. Estimating the direction and magnitude of mobility changes to restriction policies, at different pandemic phases, could offer insights into not only what worked –but for whom. In doing so, results could offer insights into future pandemic planning, shaping the implementation of swift non-pharmacological responses to mitigate spread but without amplifying health inequities.

Across countries, emerging data suggest socioeconomic differences in mobility and measures of ability to uptake, or “adhere to”, restriction policies. Huang et al. refer to the phenomena as the “*luxury of social distancing*” [6]. Among the earliest studies that used area-level mobility metrics from cell-phone data were in the United States [7, 8]. These studies found that lower-income areas and counties were associated with smaller reductions in mobility following the policies [7, 8]]. Findings were similar in the province of Ontario (Canada): higher levels of area economic dependency (a composite measure with age-structure, work-force participation, and dependency on social assistance) was associated with reduced responsiveness to restrictive policies implemented during the first year of the pandemic [9]. However, this Canadian study did not have the available data to account for expected differences in baseline mobility across socioeconomic factors; nor to compare regions in the province with and without restriction policies [9] – an important comparator used in other studies to better attribute mobility changes to specific policies [8]. Finally, composite measures of socioeconomic factors are commonly used because they capture clustering or latent features, but raw measures such as household income may provide further clarity when drawing inference. For example, pathways by which restriction policies take effect could be masked with a composite exposure measure (e.g. restriction policies would work differently for younger versus older individuals supported by social assistance). For the same reason, raw measures are commonly used to characterize patterns of SARS-CoV-2 risks [1, 10].

To address these knowledge gaps, we used area-level cell-phone based mobility and socioeconomic data in five regions in Ontario (Canada) to: (1) describe variability in the mobility response by income and by occupation in essential services following two restriction policies; and (2) estimate the extent to which these two area-level socioeconomic measures modified the mobility response, accounting for expected differences over time in the absence of restrictions using regions without restrictions as the control group.

## Methods

### Study design, setting, and population

We conducted a retrospective, observational study in accordance with the STROBE (Strengthening the Reporting of Observational Studies in Epidemiology) recommendations [11] (**S1 Checklist**). The study population comprises all census tracts in the five public health units (Toronto, Peel, Halton, York, and Durham [12], **S1 Fig**) in Ontario, Canada’s most populous province and epicenters of the SARS-CoV-2 pandemic. The five public health units make up the Greater Toronto Area (population over 7.1 million [13]) which is the largest metropolitan area in Canada. Census tracts are Statistics Canada geographic units that are only used within metropolitan areas [14]. The province of Ontario is served by a single-payer, province-wide health care system and COVID-19 related policy measures, including the restrictions, were implemented at the level of the public health unit [15]. Our analyses were conducted at the unit of census tract. We excluded census tracts with missing population size or mobility data (**S2 Fig**).

The first case of COVID-19 was reported on January 23, 2020 in the Greater Toronto Area [16]. The first province-wide restrictions were implemented on March 17, 2020 and thus, across all five public health units (**Fig 1**) [17]. Restrictions were eased over the subsequent months. The second restriction was implemented on November 23, 2020 in only two public health units (Toronto, Peel; total population 4.1 million) [18]. **S1 Table** details the nature of the restriction policies.

**Fig 1.**
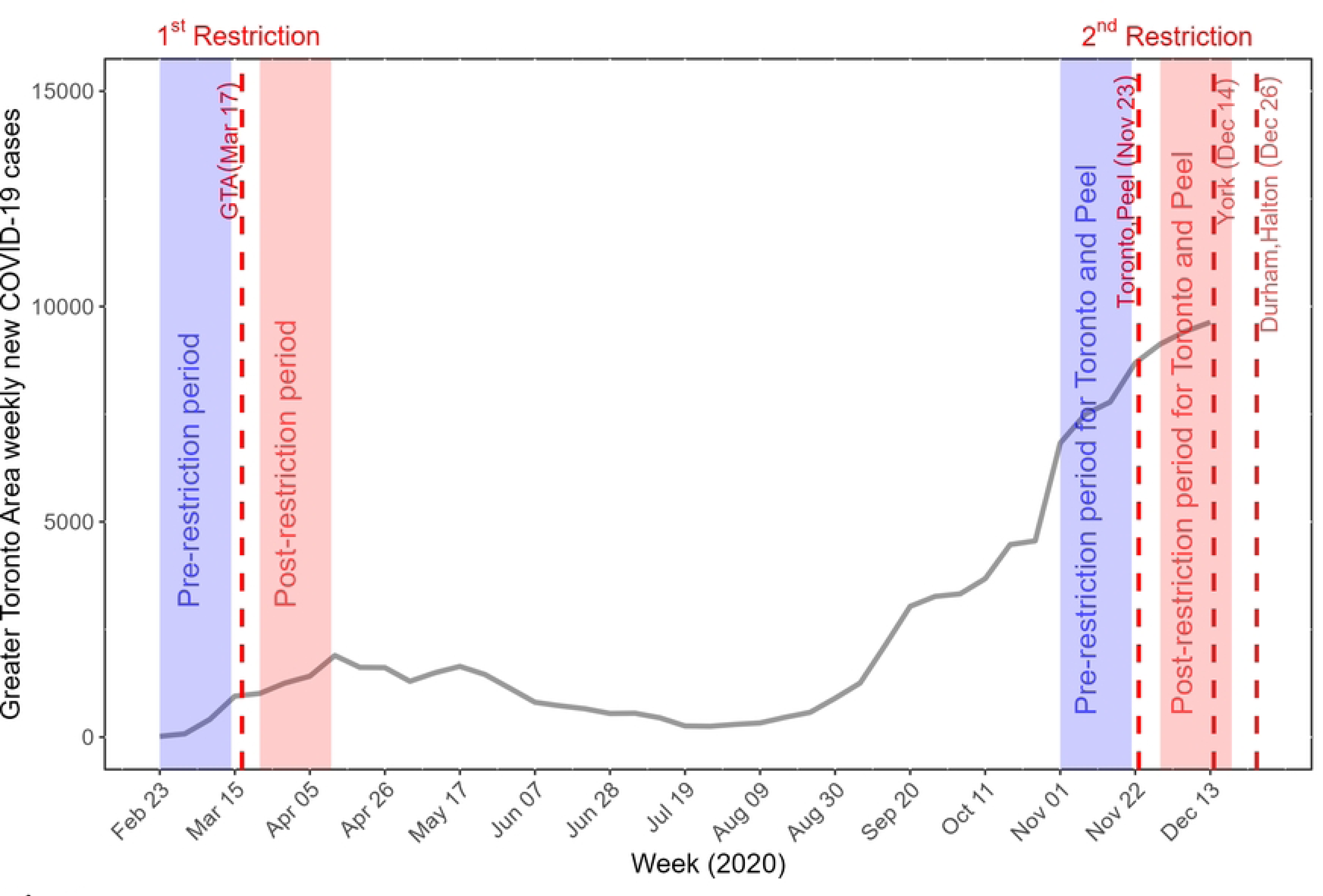
COVID-19 epidemic curve and the timing of the restrictions for the Greater Toronto Area between February 23, 2020 and December 13, 2020. The vertical dashed lines depict the timelines for two restriction policies. The first restriction was enacted March 17, 2020 across all five public health units (Toronto, Peel, Halton, York, and Durham) in the Greater Toronto Area. The second restriction enacted on November 23, 2020 in Toronto and Peel public health units and in the remaining public health units on December 14 or 26, 2020. The shaded areas in blue represent the three-week periods used in the analysis before the restrictions, and the shaded areas in red represent the three-week periods after the respective restriction was enacted. The week in which each restriction policy was enacted was excluded from the analyses. The weekly cases comprise diagnosed cases, and exclude cases among residents of long-term care homes.

### Data sources and measures

#### Mobility metric (crude and adjusted)

We used the aggregate mobility data made available by the Ontario Ministry of Health through the COVID-19 Ontario Modelling Consensus Table [19], generated by BlueDot from the data vendor Veraset (**S1 Text**). Veraset data comprises geographic position system location data across apps on different platforms with users’ consent on the use of their anonymized data, and reflects approximately 13% of the Canadian population [20]. BlueDot averaged mobility data by epidemiological week from the daily mobility metric and aggregated the data at the census-tract-level, resulting in a census-tract-level weekly average mobility metric, capturing weekly average proportion of devices that went outside their “primary location” for at least 30 minutes in a day, within each census tract. **S1 Text** details how the mobility metric was spatially and temporally aggregated.

We used the census-tract-level weekly average mobility metric (hereafter referred to as crude mobility metric) as one of our primary outcomes of interest. To account for the potential seasonal fluctuations, we generated an adjusted mobility metric in 2020 as another primary outcome of interest, by subtracting the crude mobility metric in 2019 from the crude mobility metric in 2020.

#### Pre-restriction and post-restriction periods

We defined pre- and post-restriction periods as three weeks before and after policy implementation, respectively; and excluded the week of implementation [8]. Consistent with previous studies, we selected a three-week period to capture potential lags between policy implementation and ability of individuals to respond [7]. Thus, for the first restriction, pre-restriction period captures February 23 to March 8, 2020; and post-restriction period captures March 22 to April 5, 2020. For the second restriction, pre-restriction period captures November 1 to November 15, 2020 and post-restriction period captures November 29 to December 13, 2020.

#### Socioeconomic measures

Other primary covariates of interest included two census-tract-level socioeconomic measures, generated using 2016 Canadian census data and Postal Code Conversion File Plus: income (defined by the after-tax income per-person equivalent) and the proportion of essential workers (defined based on national occupation categories and to include occupations that were not amenable to remote work: trades, transport, and equipment operation; sales and services; manufacturing and utilities; and resources, agriculture, and production [1, 10]; **S2 Table**). We excluded the health care category as it encompasses a wide range of professions with heterogeneous socioeconomic strata, and our rationale for the study was to characterize mobility patterns in the context of lower-wage essential services.

The socioeconomic measures were categorized into quintiles by ranking census tracts across the five public health units, weighted by census tract population size (**S3 Fig**). Each quintile therefore has similar population size and a different number of census tracts (**S3 Table**). For the income measure, quintile 1 refers to the highest income group. For the essential worker measure, quintile 1 refers to the lowest proportion of essential workers.

#### Other data sources

To describe the epidemic curves of COVID-19 cases, we used anonymized, person-level data on laboratory-confirmed cases (excluding residents of long-term care homes) from the provincial surveillance data (Ontario’s Case and Contact Management) between February 23, 2020 and December 13, 2020 (details in **S2 Text)** [19].

#### Ethical approval

The University of Toronto Health Sciences Research Ethics Board (protocol no. 39253) approved the study.

### Descriptive analysis: mobility change and epidemic curves

We first examined the representativeness of the mobility metric by socioeconomic measures by comparing the average census tract-level mobile device coverage across each socioeconomic measure.

To contextualize mobility changes in the context of the local epidemic, we visualized the epidemic curve alongside the crude mobility metrics overall and for each socioeconomic quintile. For the epidemic curve, we calculated weekly new lab-confirmed COVID-19 cases between February 23 and December 13, 2020. We plotted the weekly crude mobility metric for the corresponding calendar weeks in 2019 and 2020, respectively.

We examined the overall average mobility change across census tracts following each restriction by calculating the absolute difference in the mobility metrics pre- and post- each restriction. For the second restriction, we limited our analysis to Toronto and Peel only. We first calculated a weekly average mobility within each census tract (averaged over the three weeks) for each period (pre and post). We then subtracted the pre-restriction weekly average mobility from the post-restriction mobility to obtain mobility change for each census tract. We finally calculated the mean mobility change across census tracts. We repeated this for both the crude and adjusted mobility metrics.

We then examined the absolute mobility change (for crude and adjusted metrics, respectively) stratified by socioeconomic measures.

### Difference-in-differences analysis: overall impact of the second restriction on mobility

First, we conducted a difference-in-differences analysis [21] to estimate the overall influence of the second restriction on mobility in all five public health units using a mixed-effect linear model (**Model 1)** (**S3 Text**). We leveraged data from Halton, York, and Durham (regions without restriction) as the control group to account for expected mobility change over time in the absence of a restriction. Toronto and Peel which received the second restriction were treated as the ‘intervention’ group. The outcome was the adjusted mobility metric within each census tract for each epidemiological week (a total of 6 weeks data: 3 weeks pre-restriction and 3 weeks post-restriction). **S4 Fig** displays the timing of the second restriction and periods that we used for the difference-in-differences analysis. We included the following fixed effects in the model: the week index as a categorical variable reflecting each of the six weeks; the intervention indicator as a binary variable which was set to one for census tracts within ‘intervention’ group (Toronto, Peel), and set to zero for the census tracts within the control group (Halton, York, Durham). The time-dependent restriction indicator as a binary variable which was set to one if a given census tract was under restriction in a given week, and otherwise set to zero. We accounted for variances clustered at the levels of the census tract and at the public health unit by including random intercepts at the census tract and public health unit, respectively.

### Difference-in-differences analysis: impact of the second restriction on mobility stratified by socioeconomic measures

To examine the effect modification by socioeconomic measures on the relationship between the second restriction and mobility, we fitted two additional difference-in-differences mixed-effect linear models to examine the effect modification by census-tract-level income (**Model 2A**), and census-tract-level proportion of essential workers (**Model 2B**), respectively (**S3 Text)**. In each of **Model 2A** and **Model 2B**, besides fixed and random effects already shown in **Model 1**, we added socioeconomic quintiles as an additional covariate, the its interactions with every other covariates in **Model 1**. We conducted the two-way analysis of variance to test whether there was evidence of effect modification on the restriction effect by socioeconomic measures. We herein refer to a census tract as a neighborhood in the following sections.

## Results

The five public health units in the current study include 1,261 census tracts, with population sizes ranging from 484 to 23,401. We excluded 7 census tracts with missing population size data and another 14 census tracts with missing mobility data (**S2 Fig**). Of 1240 census tracts included, the mean device coverage was lower in neighborhoods with the lower income (**S4 Table**).

### Epidemic curves

Following the first restriction, the epidemic rapidly diverged along socioeconomic quintiles such that neighborhoods with lower-income and neighborhoods with higher proportion essential workers experienced higher number of COVID-19 cases (**Fig 2A-B**). In August 2020, the number of weekly new lab-confirmed cases were small across all socioeconomic quintiles before diverging by mid-September 2020, with the highest number of cases in lowest-income neighborhoods and in neighborhoods with highest proportion of essential workers. The number of cases remained divergent along socioeconomic quintiles prior to and following the second restriction.

**Fig 2.**
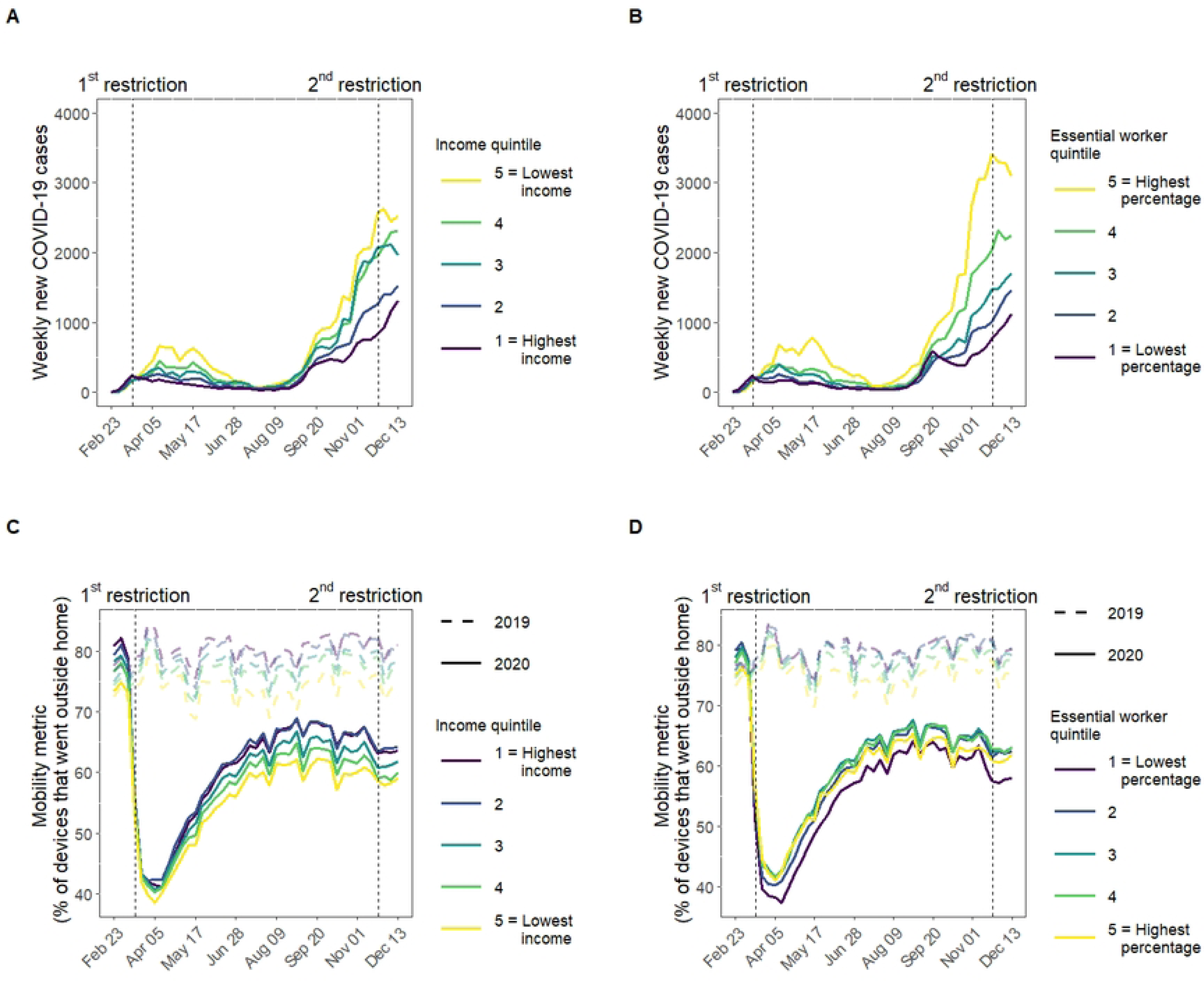
Epidemic curves and mobility change stratified by neighborhood-level income and essential worker quintiles in the Greater Toronto Area, Canada (February 23, 2020 to December 13, 2020). Panel A and B describe weekly new lab-confirmed COVID-19 cases (excluding residents of long-term care homes) by neighborhood-level income quintile (Panel A) and proportion of the working population engaged in essential services (Panel B). Panel C and D depict the weekly crude mobility metric by neighborhood-level income quintile (Panel C) and proportion essential workers (Panel D). The horizontal dashed lines in Panel C and D represent 2019 mobility as pre-pandemic mobility reference, while the solid lines represent 2020 mobility data. The vertical dashed lines depict the two COVID-19 restriction policies under examination: the first restriction enacted March 17, 2020 across all five public health units (Toronto, Peel, Halton, York, and Durham) in the Greater Toronto Area; and the second restriction enacted November 23, 2020 in Toronto and Peel public health units within the Greater Toronto Area.

Income reflects the per-person equivalent income in the household. Essential services include: trades, transport, and equipment operation; sales and services; manufacturing and utilities; and resources, agriculture, and production. Neighborhood level is defined at the level of the census tract. Quintiles are weighted by neighborhood-level population.

### Descriptive analysis: mobility change following the first restriction

Higher-income neighborhoods consistently demonstrated greater mobility throughout 2019 and 2020 (**Fig 2C**). In 2019, the crude mobility ranged between 80-90% in the highest-income neighborhoods and 70-75% in the lowest-income neighborhoods. In early 2020 (pre-restriction period), the crude mobility was higher in higher-income neighborhoods (e.g., 80.6% in the highest-income and 73.7% in the lowest-income neighborhoods) (**Table 1**).

**Table 1.**
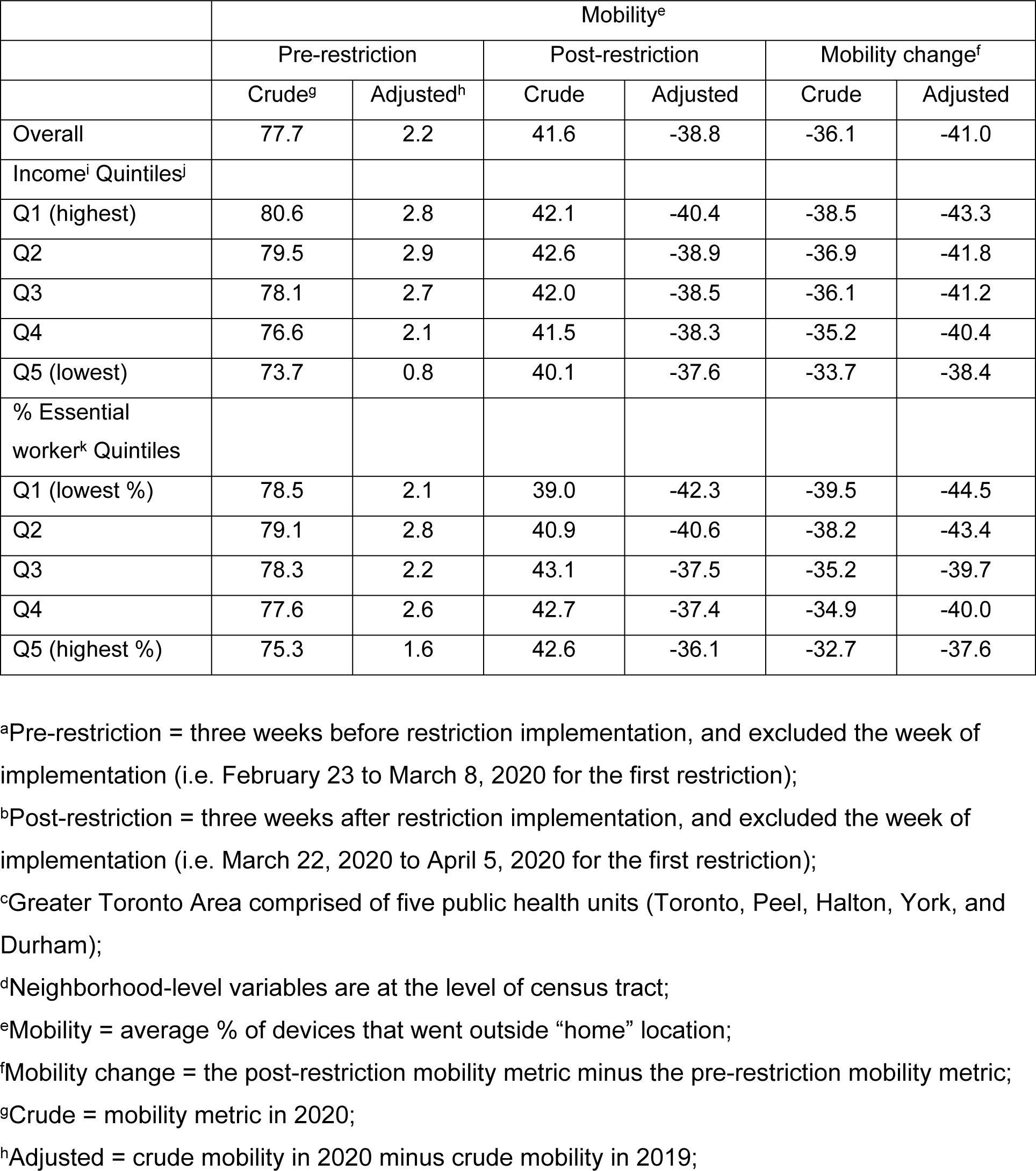
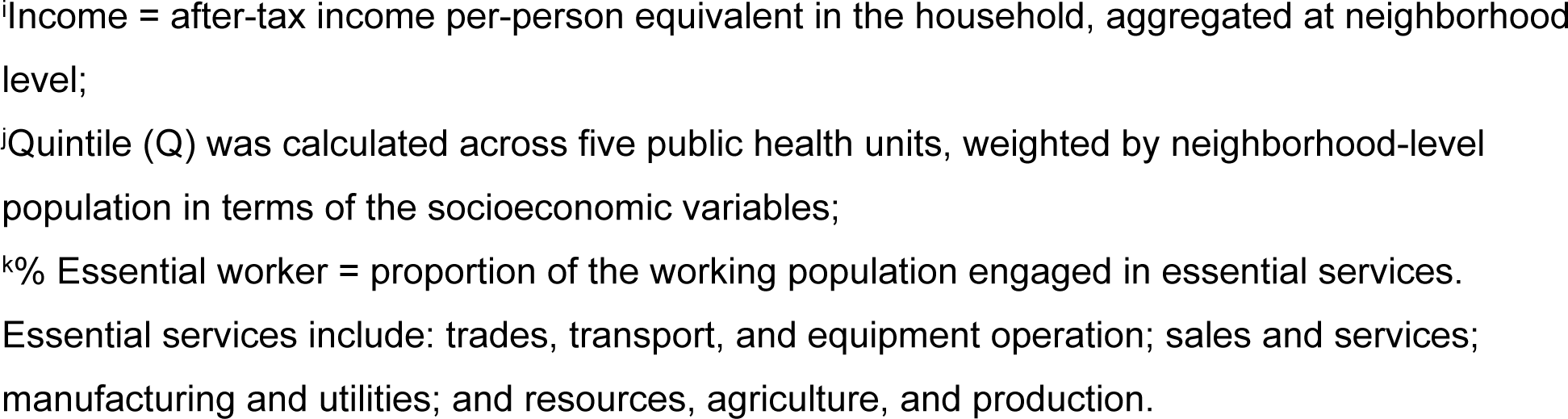
Mobility metric of pre-restriction^a^ and post-restriction^b^ periods for the first restriction in Greater Toronto Area^c^ stratified by neighborhood-level^d^ socioeconomic measures.

|  | Mobility<sup>e</sup> |  |  |  |  |  |
| --- | --- | --- | --- | --- | --- | --- |
|  | Pre-restriction |  | Post-restriction |  | Mobility change<sup>f</sup> |  |
|  | Crude<sup>g</sup> | Adjusted<sup>h</sup> | Crude | Adjusted | Crude | Adjusted |
| Overall | 77.7 | 2.2 | 41.6 | -38.8 | -36.1 | -41.0 |
| Income<sup>i</sup> Quintiles<sup>j</sup> |  |  |  |  |  |  |
| Q1 (highest) | 80.6 | 2.8 | 42.1 | -40.4 | -38.5 | -43.3 |
| Q2 | 79.5 | 2.9 | 42.6 | -38.9 | -36.9 | -41.8 |
| Q3 | 78.1 | 2.7 | 42.0 | -38.5 | -36.1 | -41.2 |
| Q4 | 76.6 | 2.1 | 41.5 | -38.3 | -35.2 | -40.4 |
| Q5 (lowest) | 73.7 | 0.8 | 40.1 | -37.6 | -33.7 | -38.4 |
| % Essential worker<sup>k</sup> Quintiles |  |  |  |  |  |  |
| Q1 (lowest %) | 78.5 | 2.1 | 39.0 | -42.3 | -39.5 | -44.5 |
| Q2 | 79.1 | 2.8 | 40.9 | -40.6 | -38.2 | -43.4 |
| Q3 | 78.3 | 2.2 | 43.1 | -37.5 | -35.2 | -39.7 |
| Q4 | 77.6 | 2.6 | 42.7 | -37.4 | -34.9 | -40.0 |
| Q5 (highest %) | 75.3 | 1.6 | 42.6 | -36.1 | -32.7 | -37.6 |

<sup>a</sup>Pre-restriction = three weeks before restriction implementation, and excluded the week of implementation (i.e. February 23 to March 8, 2020 for the first restriction);
<sup>b</sup>Post-restriction = three weeks after restriction implementation, and excluded the week of implementation (i.e. March 22, 2020 to April 5, 2020 for the first restriction);
<sup>c</sup>Greater Toronto Area comprised of five public health units (Toronto, Peel, Halton, York, and Durham);
<sup>d</sup>Neighborhood-level variables are at the level of census tract;
<sup>e</sup>Mobility = average % of devices that went outside “home” location;
<sup>f</sup>Mobility change = the post-restriction mobility metric minus the pre-restriction mobility metric;
<sup>g</sup>Crude = mobility metric in 2020;
<sup>h</sup>Adjusted = crude mobility in 2020 minus crude mobility in 2019;
<sup>i</sup>Income = after-tax income per-person equivalent in the household, aggregated at neighborhood level;
<sup>j</sup>Quintile (Q) was calculated across five public health units, weighted by neighborhood-level population in terms of the socioeconomic variables;
<sup>k</sup>% Essential worker = proportion of the working population engaged in essential services. Essential services include: trades, transport, and equipment operation; sales and services; manufacturing and utilities; and resources, agriculture, and production.

The overall crude mobility declined sharply at the onset of the first restriction from 77.7% to 41.6% (**S5 Fig**; **Table 1; S6A-B Fig**). All income quintiles experienced a decline in mobility following the restriction; however the higher-income neighborhoods experienced larger reductions than lower-income neighborhoods (**Fig 2C**). In the post-restriction period, the crude mobility was similar across income quintiles (ranging from 40.1% to 42.6%). After accounting for seasonal fluctuations, the adjusted mobility change (reduction) following the first restriction was 43.3% in the highest-income and 38.4% in the lowest-income neighborhoods with a dose-response pattern (**Table 1**).

The mobility pattern for proportion essential workers mimicked (inversely) the pattern with income (**Fig 2C-D**). The adjusted mobility reduction was 44.5% and 37.6% in neighborhoods with the lowest and highest proportion essential workers, respectively.

### Descriptive analysis: mobility change following the second restriction

After the large reduction following the first restriction, mobility resumed steadily since early April and reached a plateau in July through November 2020 across the five public health units (**S5 Fig**). During this plateau, mobility remained the lowest in the lowest-income neighborhoods (**Fig 2C**).

Following the second restriction in Toronto and Peel, the overall crude mobility experienced a small reduction (-2.8%) (**Table 2**; **S6C-D Fig**). After accounting for seasonal fluctuations, the adjusted mobility reduction in these two public health units were small (-0.8%) (**Table 2**). The adjusted mobility reduction was larger in the higher-income neighborhoods (-2.7% in the highest-income neighborhoods and -0.1% in the lowest-income neighborhoods). The patterns in mobility change by essential worker quintiles were similar, with the largest reduction in adjusted mobility observed in the neighborhoods with lowest proportion essential workers (-2.3%).

**Table 2.**
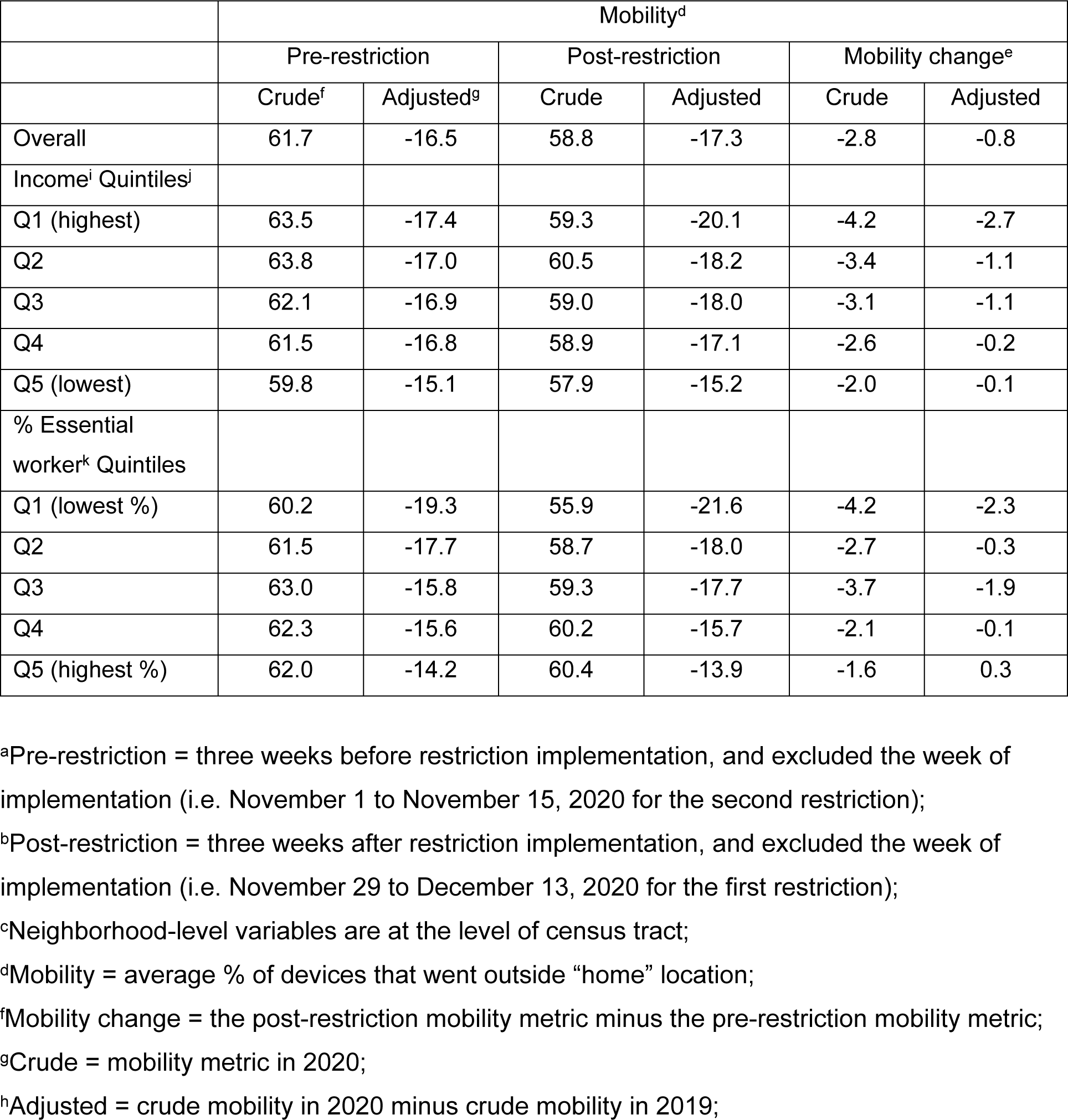
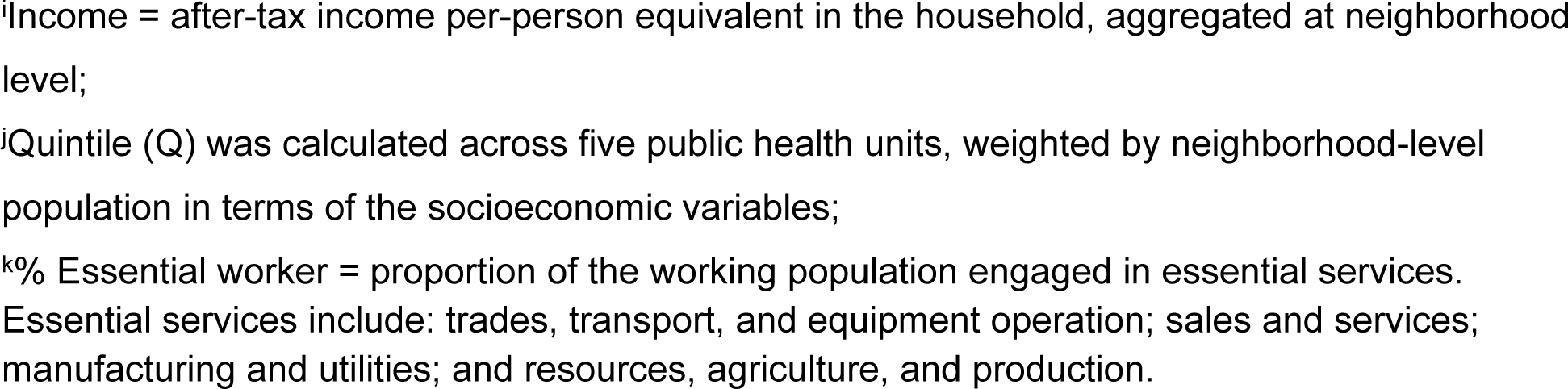
Mobility metric of pre-restriction^a^ and post-restriction^b^ periods for the second restriction in two public health units (Toronto, Peel) stratified by neighborhood-level^c^ socioeconomic measures.

|  | Mobility<sup>d</sup> |  |  |  |  |  |
| --- | --- | --- | --- | --- | --- | --- |
|  | Pre-restriction |  | Post-restriction |  | Mobility change<sup>e</sup> |  |
|  | Crude<sup>f</sup> | Adjusted<sup>g</sup> | Crude | Adjusted | Crude | Adjusted |
| Overall | 61.7 | -16.5 | 58.8 | -17.3 | -2.8 | -0.8 |
| Income<sup>i</sup> Quintiles<sup>j</sup> |  |  |  |  |  |  |
| Q1 (highest) | 63.5 | -17.4 | 59.3 | -20.1 | -4.2 | -2.7 |
| Q2 | 63.8 | -17.0 | 60.5 | -18.2 | -3.4 | -1.1 |
| Q3 | 62.1 | -16.9 | 59.0 | -18.0 | -3.1 | -1.1 |
| Q4 | 61.5 | -16.8 | 58.9 | -17.1 | -2.6 | -0.2 |
| Q5 (lowest) | 59.8 | -15.1 | 57.9 | -15.2 | -2.0 | -0.1 |
| % Essential worker<sup>k</sup> Quintiles |  |  |  |  |  |  |
| Q1 (lowest %) | 60.2 | -19.3 | 55.9 | -21.6 | -4.2 | -2.3 |
| Q2 | 61.5 | -17.7 | 58.7 | -18.0 | -2.7 | -0.3 |
| Q3 | 63.0 | -15.8 | 59.3 | -17.7 | -3.7 | -1.9 |
| Q4 | 62.3 | -15.6 | 60.2 | -15.7 | -2.1 | -0.1 |
| Q5 (highest %) | 62.0 | -14.2 | 60.4 | -13.9 | -1.6 | 0.3 |

<sup>a</sup>Pre-restriction = three weeks before restriction implementation, and excluded the week of implementation (i.e. November 1 to November 15, 2020 for the second restriction);
<sup>b</sup>Post-restriction = three weeks after restriction implementation, and excluded the week of implementation (i.e. November 29 to December 13, 2020 for the first restriction);
<sup>c</sup>Neighborhood-level variables are at the level of census tract;
<sup>d</sup>Mobility = average % of devices that went outside “home” location;
<sup>f</sup>Mobility change = the post-restriction mobility metric minus the pre-restriction mobility metric;
<sup>g</sup>Crude = mobility metric in 2020;
<sup>h</sup>Adjusted = crude mobility in 2020 minus crude mobility in 2019;
<sup>l</sup>Income = after-tax income per-person equivalent in the household, aggregated at neighborhood level;
<sup>j</sup>Quintile (Q) was calculated across five public health units, weighted by neighborhood-level population in terms of the socioeconomic variables;
<sup>k</sup>% Essential worker = proportion of the working population engaged in essential services. Essential services include: trades, transport, and equipment operation; sales and services; manufacturing and utilities; and resources, agriculture, and production.

However, we did not observe a clear dose-response pattern across other essential worker quintiles (**Table 2**).

### Difference-in-differences analysis: mobility change following the second restriction

The difference-in-difference analyses in Model 1 showed that after accounting for seasonal fluctuation, and expected changes over time in the absence of the restriction, the second restriction was associated with a small overall reduction in the adjusted mobility: -0.96% (95% confidence interval (CI): (-1.53; -0.38)).

There was effect modification by income quintiles (*p* < 0.05 in **Model 2A**) and by essential worker quintiles (*p* < 0.05 in **Model 2B**) on the relationship between the second restriction and adjusted mobility. Neighborhoods with lower income and higher proportion essential workers dampened the magnitude of adjusted mobility change, consistent with the descriptive findings. However, given the small magnitude of the associations, a consistent dose-response pattern was not observed (**Fig 3**; **S5 Table**).

**Fig 3.**
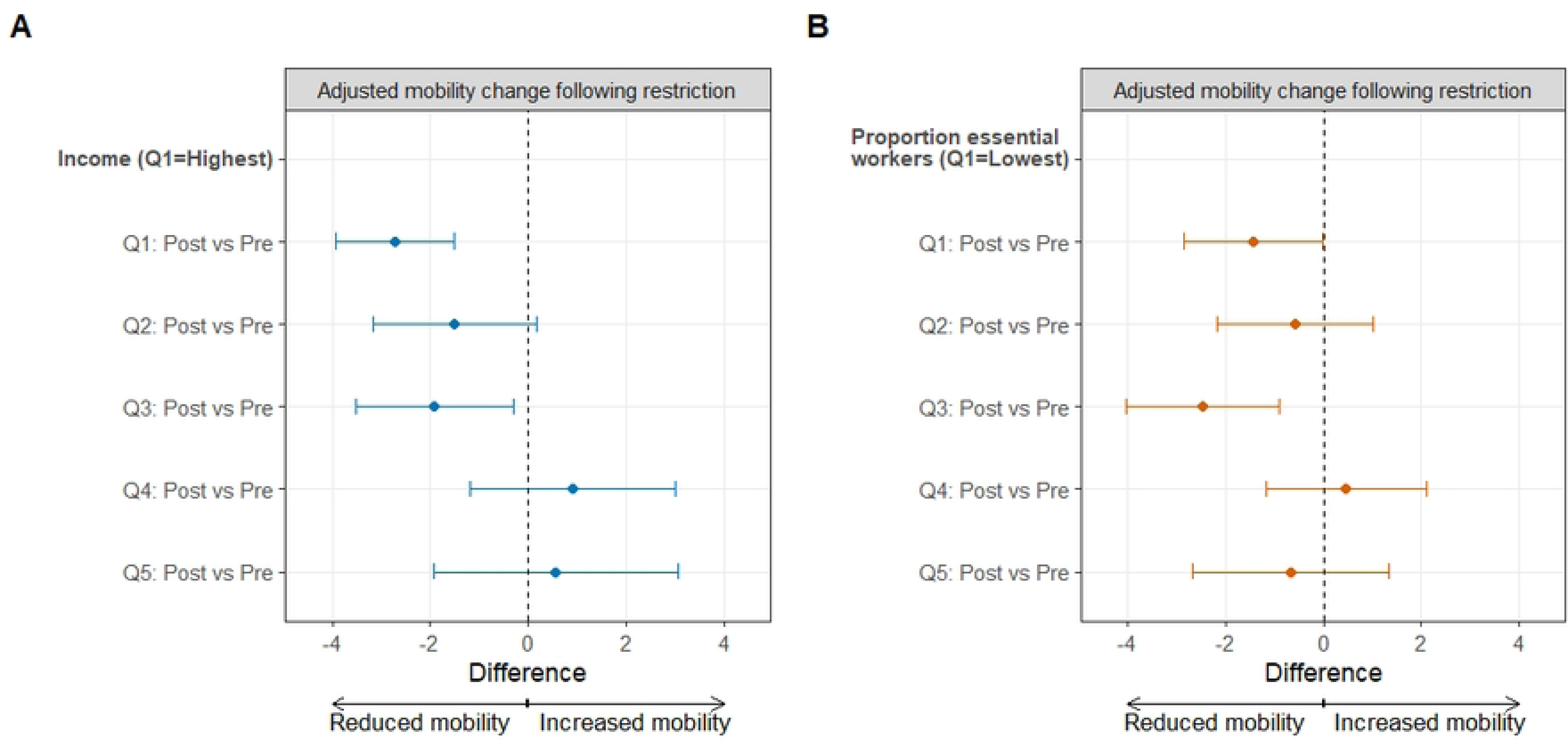
Adjusted mobility change following the second restriction by neighborhood-level socioeconomic measures in the Greater Toronto Area. Panel A shows the estimates of the adjusted mobility change following the second set of restrictions by income quintiles. Panel B shows the estimates of the adjusted mobility following the second set of restrictions by essential worker quintiles. The error bars represent the 95% confidence interval. The Greater Toronto Area comprised of five public health units (Toronto, Peel, Halton, York, and Durham). Income reflects the per-person equivalent income in the household. Essential services include: trades, transport, and equipment operation; sales and services; manufacturing and utilities; and resources, agriculture, and production. Neighborhood level is defined at the level of the census tract. Quintiles (Q) are weighted by neighborhood-level population.

## Discussion

Using an ecological study of neighborhood-level mobility measures in Ontario, Canada during 2020, we found that the first COVID-19 restriction led to large (30-40%) reductions in crude mobility. Reduction in mobility following the first restriction was largest in the highest-income neighborhoods and neighborhoods with the fewest essential workers. In contrast, a very small (<1%) reduction in mobility could be attributed to the second restriction. However, there was still evidence of effect modification by income and proportion essential workers.

The large effect of the first restriction followed by a much smaller effect of the second restriction is consistent with findings that demonstrate a similar temporal saturation effect of public health measures over subsequent COVID-19 waves [22, 23]. The first restriction occurred during a time of large uncertainty. Then, even as some elements of the restrictions were lifted, there remained an overall – and relatively stable – level of mobility that was a third lower than levels in 2019. This “new normal” in 2020 reflected a shift from in-person to online shopping, education, socializing, and especially working – via occupations amenable to remote work and across businesses that were able to shift to largely remote production [24]. This time-period was also marked by a 4.8% loss of employment in the province [25]. When the second restriction was implemented to try and mitigate rising cases of COVID-19 during Ontario’s second epidemic wave, it had very little effect on mobility in the context of the “new normal” that had been established. For example, workplaces that could transition to remote work had already done so. A similar saturation effect of increasing levels of stringency of COVID-19 measures on cases was observed across multiple provinces in Canada, with little effect of stringency measures during the second COVID-19 waves that occurred between August and December 2020 [23].

Taken together, the findings highlight the importance of anticipating how the timing of restrictions and the wider context of what is already in place could influence the impact of the restriction.

The mobility changes to the first and second restriction varied by income and by occupation, and suggest a saturation effect of restrictions by socioeconomic characteristics. The mobility change was lower in lower-income neighborhoods, and in those with higher proportion essential workers. This finding was consistent with prior studies from the United States, Italy, and Canada that had examined income [7, 9, 26, 27], and from survey data in Canada that similarly reported lower levels of ability to “shelter in place” among lower-income households [28]. Our study separated the analyses of income from that of occupation, and confirmed a similar pattern with occupation to that of income. That is, as hypothesized, neighborhoods with higher proportion essential workers experienced a smaller mobility change to the restrictions. This smaller mobility change occurred against a pre-pandemic background wherein neighborhoods with more essential workers, and lower income, already had lower levels of mobility. The 2019 mobility patterns capture non-occupational travel, including travelling outside the home for shopping and recreation and suggests that people living in lower-income have less access to the latter at baseline [29]. This baseline difference means that individuals in lower-income neighborhoods and in neighborhoods with higher proportion essential workers may have already had fewer non-occupational activities to limit when restrictions were put into place. Indeed, mobility data from the US suggest that although people living in lower-income neighborhoods were more likely to work outside the home during the pandemic, they were less likely to have access to and visit recreational venues such as parks [29]. Such findings [29] may help also explain why despite variability in the mobility change by income and occupation, the magnitude of difference between socioeconomic levels was small – especially after the second restriction. That is, in addition to baseline lower levels of mobility (presumably from non-occupational travel), individuals may have reduced mobility in other ways to make up for having to work onsite.

Although neighborhood-level income and proportion essential workers are somewhat correlated [10], they represent related but distinct social determinants of mobility. For example, the pathways by which the restriction acts (i.e. its “mechanisms of action”) on mobility across income levels is complex and, in addition to occupation-related travel, can also be related to household size (larger households offer more opportunities for cellular signals of travelling outside the home), caregiver roles (intergenerational household dependents who may require healthcare visits), access to services (e.g. grocery chains for home delivery) and access to household greenspace (backyards). Taken together, baseline (pre-pandemic) variability in mobility combined with differences in one’s ability to transition to remote work by income and by occupation highlight the saturation effect of restrictions by socioeconomic characteristics [8].

This socioeconomic saturation effect stems from the extent to which a policy or intervention is designed to reach and work across subsets of the population.

Our study was not designed to quantify the impact of differential mobility change on differential rates of SARS-CoV-2 cases. However, the small magnitude of difference in mobility reduction by socioeconomic characteristics, especially after the second restriction, suggests that differential mobility may be insufficient to explain the difference in SARS-CoV-2 cases by socioeconomic characteristics in the region. The implication would be residual risks of exposures and transmission that were not addressed by the restrictions in 2020, including onsite workplace exposures intersecting with high-density and multigenerational households [1, 30]. The findings surrounding extant but small differences in mobility change alongside large disparities in SARS-CoV-2 cases suggests the need for additional, tailored approaches to address residual risks, such as outreach testing and comprehensive isolation support, vaccination, and changes in policies such as paid sick leave [31, 32].

Our study has several limitations. First, our outcome of interest was a mobility metric, which served as a proxy for contacts between people. The metric (proportion of devices that travelled outside “primary location”) undervalues contacts within households (an important space where transmission occurs) and simultaneously overvalues what happens outside the home (i.e. leaving home but not coming into close contact with others). The restriction was only designed to reduce contacts outside the home. If our metric is a proxy for contacts outside the home, then our study may have underestimated the magnitude of variability in mobility reduction by socioeconomic levels. For example, our metric for mobility may be more likely to capture travel for onsite work among individuals living in lower-income neighborhoods as compared to individuals living in higher-income neighborhoods, where our metric may be more likely to capture mobility for non-occupational travel. If that were the case, and mobility for onsite work is related to more workplace contacts than travel outside the home for other reasons, then there may exist an even larger socioeconomic difference in contacts outside the home than our analyses would suggest. Data are emerging from self-reported data on contacts between people and their correlation with area-level mobility metrics derived from cell-phones. [33]; these correlations, particularly if they differ by socioeconomic levels, offer an opportunity for potential bias-adjustment or correction factors when using cell-phone mobility metrics to evaluate the impact of restriction policies on mobility changes, or downstream outcomes such as SARS-CoV- 2 cases [34]. A second limitation that could underestimate variability in our outcome of interest stemps undersampling of mobility measures in lower-income neighborhoods. We also restricted our study to a mobility metric commonly used across prior research and public health teams [35]. Future work could benefit from examining other mobility measures, such as proportion of time away from home. The magnitude of change in contact rates (and thus, its proxy – the mobility metric) that is needed to reduce transmission depends on the underlying transmission potential at the time. That is, how much of a reduction is needed is different at different stages of an epidemic and in different contexts of underlying risks. A smaller reduction in contacts may be needed among high-income households with less crowding and lack of occupational SARS-CoV-2 risks. Finally, we were restricted to neighborhood-level aggregated information. This means that both the socioeconomic variables and the mobility metrics were averaged over a population and may not reflect individual-level patterns. However, individual-level survey data suggest the ecological findings from our study are consistent with self-reported data on income and mobility in response to public health restrictions in Canada [28].

In summary, restrictions used in the COVID-19 public health response in 2020 demonstrated a temporal saturation effect over subsequent waves. Restrictions also demonstrated a saturation effect by income and occupation. However, the magnitude of difference between socioeconomic levels was small – especially after the second restriction. At the same time, there was a consistent and large difference in SARS-CoV-2 cases by socioeconomic characteristics which suggest residual transmission risks along socioeconomic margins. Findings highlight the need for additional approaches to reduce health inequities at the intersections of income and occupation when addressing large epidemics of novel and resurging respiratory pathogens.

## Data Availability

Socioeconomic variables were generated from of the 2016 census data (https://www12.statcan.gc.ca/census-recensement/2016/dp-pd/index-Eng.cfm) and Postal Code Conversion File Plus (https://www12.statcan.gc.ca/census-recensement/2016/dp-pd/index-Eng.cfm) and are publicly available from Statistics Canada. The Ontario’s Case and Contact Management data on the laboratory-confirmed COVID-19 cases and mobility data used in this study are not publicly available but can be requested the Government of Ontario Ministry of Health (https://www.ontario.ca/feedback/contact-us?id=532365&nid=138269).

## Acknowledgements

We thank Dr. Alexander Watts (BlueDot) for insights and support on the mobility data. We thank Samantha Lo (Unity Health Toronto) and Kristy Yiu (Unity Health Toronto) for support with research coordination. SM is supported by Tier 2 Canada Research Chair (CRC) in Mathematical Modeling and Program Science (CRC no. 950-232643). MMG is supported by a Tier 2 Canada Research Chair in Population Health Modeling.

## Supporting Information captions

**S1 Text. Definition of the mobility metric: % of devices that went outside home location**

**S2 Text. Source of COVID-19 person-level data**

**S3 Text. Model equations for difference-in-differences analysis with mixed-effect models for the 2^nd^ restriction policy**

**S1 Table. Measures for 1^st^ restriction and 2^nd^ restriction of COVID-19 in Ontario**

**S2 Table. Socioeconomic variables from Statistics Canada 2016 Census of Population**

**S3 Table. Neighborhood-level socioeconomic characteristics across 1254 census tracts in the Greater Toronto Area.**

**S4 Table. Device penetration in 1240 census tracts within the Greater Toronto Area stratified by neighborhood-level socioeconomic measures during the pre-restriction and post-restriction periods related to the two restrictions examined in the study.**

**S5 Table. Difference-in-differences analysis of the second restriction with mixed-effect modeling in Greater Toronto Area by area-level socioeconomic measures.**

**S1 Fig. Five public health units consist of census tracts within the Greater Toronto Area.**

**S2 Fig. Flow diagram for data process and for descriptive analyses and difference-in-differences analyses by mixed-effect model.**

**S3 Fig. The geographic distribution of income quintiles and essential worker quintiles at the census tract level in five public health units within the Greater Toronto Area.**

**S4 Fig. Epidemic curve and the restriction policy timing for the intervention group (Toronto and Peel) and the control group (York, Durham and Halton) in Greater Toronto Area**.

**S5 Fig. Mobility trajectories for 2019 and 2020 with the timing of two restrictions in the Greater Toronto Area.**

**S6 Fig. Maps for the three-week average mobility before and after 1^st^ restriction in five public health units within the Greater Toronto Area, and three-week average mobility before and after 2^nd^ restriction in Toronto and Peel public health units.**

**S1 Checklist. STROBE Statement**

## References

1. Xia Y, Ma H, Moloney G, García HAV, Sirski M, Janjua NZ, et al. Geographic concentration of SARS-CoV-2 cases by social determinants of health in metropolitan areas in Canada: a cross-sectional study. CMAJ. 2022;194(6):E195–E204.

2. Mishra S, Kwong JC, Chan AK, Baral SD. Understanding heterogeneity to inform the public health response to COVID-19 in Canada. Cmaj. 2020;192(25):E684–E5.

3. Gonzalez CJ, Aristega Almeida B, Corpuz GS, Mora HA, Aladesuru O, Shapiro MF, et al. Challenges with social distancing during the COVID-19 pandemic among Hispanics in New York City: A qualitative study. BMC Public Health. 2021;21(1):1–8.

4. Messacar D, Morissette R, Deng Z. Inequality in the Feasibility of Working from Home during and after COVID-19. Statistics Canada= Statistique Canada; 2020.

5. Rao A, Ma H, Moloney G, Kwong JC, Jüni P, Sander B, et al. A disproportionate epidemic: COVID-19 cases and deaths among essential workers in Toronto, Canada. Annals of epidemiology. 2021;63:63–7.

6. Huang X, Li Z, Jiang Y, Ye X, Deng C, Zhang J, et al. The characteristics of multi-source mobility datasets and how they reveal the luxury nature of social distancing in the US during the COVID-19 pandemic. International Journal of Digital Earth. 2021;14(4):424–42.

7. Weill JA, Stigler M, Deschenes O, Springborn MR. Social distancing responses to COVID-19 emergency declarations strongly differentiated by income. Proceedings of the national academy of sciences. 2020;117(33):19658–60.

8. Jay J, Bor J, Nsoesie EO, Lipson SK, Jones DK, Galea S, et al. Neighbourhood income and physical distancing during the COVID-19 pandemic in the United States. Nature Human Behaviour. 2020;4. doi: 10.1038/s41562-020-00998-2.

9. Long JA, Ren C. Associations between mobility and socio-economic indicators vary across the timeline of the Covid-19 pandemic. Computers, environment and urban systems. 2022;91:101710.

10. Mishra S, Ma H, Moloney G, Yiu KCY, Darvin D, Landsman D, et al. Increasing concentration of COVID-19 by socioeconomic determinants and geography in Toronto, Canada: an observational study. Annals of Epidemiology. 2021. doi: 10.1016/j.annepidem.2021.07.007.

11. Von Elm E, Altman DG, Egger M, Pocock SJ, Gøtzsche PC, Vandenbroucke JP. The Strengthening the Reporting of Observational Studies in Epidemiology (STROBE) statement: guidelines for reporting observational studies. The Lancet. 2007;370(9596):1453-7.

12. Public Health Units: Association of Local Public Health Agencies; [cited 2023 19 March]. Available from: https://www.alphaweb.org/page/PHU.

13. Ontario population projections: Government of Ontario; 2021 [cited 2022 August 11]. Available from: https://www.ontario.ca/page/ontario-population-projections.

14. Census Profile, 2016 Census: Statistics Canada; 2017 [cited 2022 July 12]. Available from: https://www12.statcan.gc.ca/census-recensement/2016/dp-pd/prof/index.cfm?Lang=E.

15. Ontario Public Health System Public Health Onatrio [cited 2022 July 12]. Available from: https://www.publichealthontario.ca/en/About/News/2020/Ontario-Public-Health-System#:~:text=Public%20Health%20Ontario,-Public%20Health%20Ontario&text=Our%20expertise%20spans%20the%20following,prevention%2C%20infectious%20disease%20and%20microbiology.

16. Urrutia D, Manetti E, Williamson M, Lequy E. Overview of Canada’s answer to the COVID-19 pandemic’s first wave (January–April 2020). International Journal of Environmental Research and Public Health. 2021;18(13):7131.

17. Nielsen K. A timeline of COVID-19 in Ontario. Global News. April 24, 2020.

18. Ontario declares second provincial emergency to address COVID-19 crisis and save lives: Government of Ontario; 2021 [cited 2022 July 15]. Available from: https://news.ontario.ca/en/release/59922/ontario-declares-second-provincial-emergency-to-address-covid-19-crisis-and-save-lives.

19. Hillmer MP, Feng P, McLaughlin JR, Murty VK, Sander B, Greenberg A, et al. Ontario’s COVID-19 Modelling Consensus Table: mobilizing scientific expertise to support pandemic response. Canadian Journal of Public Health. 2021;112(5):799–806.

20. COVID-19 Risk Assessment: Social Distancing in Canada, prepared by BlueDot, March 27th, 2020. 2020.

21. Wing C, Simon K, Bello-Gomez RA. Designing difference in difference studies: best practices for public health policy research. Annual review of public health. 2018;39:453–69.

22. Kim J, Kwan M-P. The impact of the COVID-19 pandemic on people’s mobility: A longitudinal study of the US from March to September of 2020. Journal of Transport Geography. 2021;93:103039.

23. Vickers DM, Baral S, Mishra S, Kwong JC, Sundaram M, Katz A, et al. Stringency of containment and closures on the growth of SARS-CoV-2 in Canada prior to accelerated vaccine roll-out. International Journal of Infectious Diseases. 2022;118:73–82.

24. Clarke S. Working from home during the Covid-19 pandemic: How rates in Canada and the United States compare: Statistics Canada= Statistique Canada; 2022.

25. COVID-19 Pandemic Causes Record Job Loss. Financial Accountability Office of Ontario, 2021.

26. Bonaccorsi G, Pierri F, Cinelli M, Flori A, Galeazzi A, Porcelli F, et al. Economic and social consequences of human mobility restrictions under COVID-19. Proceedings of the National Academy of Sciences. 2020;117(27):15530–5.

27. Marwah A, Feldman J, Moineddin R, Thomas A. Population mobility and socioeconomic indicators in California, USA and Ontario, Canada during the COVID-19 pandemic. International Journal of Infectious Diseases. 2022;116:S25-S6.

28. Lavoie KL, Gosselin-Boucher V, Stojanovic J, Voisard B, Szczepanik G, Boyle JA, et al. Determinants of adherence to COVID-19 preventive behaviours in Canada: Results from the iCARE Study. MedRxiv. 2021:2021.06. 09.21258634.

29. Jay J, Heykoop F, Hwang L, Courtepatte A, de Jong J, Kondo M. Use of smartphone mobility data to analyze city park visits during the COVID-19 pandemic. Landscape and Urban Planning. 2022;228:104554.

30. Ma H, Yiu KC, Baral SD, Fahim C, Moloney G, Darvin D, et al. COVID-19 Cases Among Congregate Care Facility Staff by Neighborhood of Residence and Social and Structural Determinants: Observational Study. JMIR Public Health and Surveillance. 2022;8(10):e34927.

31. Kiran T, Eissa A, Mangin D. Brief on Primary Care Part 1: The roles of primary care clinicians and practices in the first two years of the COVID-19 pandemic in Ontario. Science Briefs of the Ontario COVID-19 Science Advisory Table. 2022; 3 (67). 2022.

32. Adam Nagy AMC, Margaret Bourdeaux. Organizing, Budgeting, and Implementing Wraparound Services for People in Quarantine and Isolation Harvard University: Berkman Klein Center for Internet & Society; 2021 [cited 2023 October 18, 2023]. Available from: https://cyber.harvard.edu/story/2021-03/organizing-budgeting-and-implementing-wraparound-services-people-quarantine-and.

33. Barrios JM, Benmelech E, Hochberg YV, Sapienza P, Zingales L. Civic capital and social distancing during the Covid-19 pandemic☆. Journal of public economics. 2021;193:104310.

34. Tomori DV, Rübsamen N, Berger T, Scholz S, Walde J, Wittenberg I, et al. Individual social contact data and population mobility data as early markers of SARS-CoV-2 transmission dynamics during the first wave in Germany—an analysis based on the COVIMOD study. BMC medicine. 2021;19:1–13.

35. Brown KA, Soucy J-PR, Buchan SA, Sturrock SL, Berry I, Stall NM, et al. The mobility gap: estimating mobility thresholds required to control SARS-CoV-2 in Canada. CMAJ : Canadian Medical Association journal = journal de l’Association medicale canadienne. 2021;193. doi: 10.1503/cmaj.210132. PubMed PMID: 33827852.

